# Joinpoint Regression to Determine the Impact of COVID-19 on Mortality in Europe: A Longitudinal Analysis From 2000 to 2020 in 27 Countries

**DOI:** 10.1101/2022.01.19.22269576

**Authors:** Alessandro Rovetta

**Affiliations:** R&C Research

## Abstract

The novel coronavirus disease 2019 (COVID-19) represented the most extensive health emergency in human history. However, to date, there is still a lot of uncertainty about the exact death toll the pandemic has claimed. In particular, the number of official deaths could be vastly underestimated. Despite this, many conspirationists speculate that COVID-19 is not a dangerous disease. Therefore, in this manuscript, we use joinpoint regression analysis to estimate the impact of COVID-19 in 27 European countries by comparing annual mortality trends from 2000 to 2020. Furthermore, we provide accessible evidence even for a non-expert audience. Siegel (A1) and Holm-Bonferroni (A2) approaches were employed to assess the significance of the results separately. In conclusion, these results estimate that COVID-19 increased the overall mortality in Europe by 10% (A1: P < .001, A2: Adjusted P = .001). In 16 out of 27 countries (59.3%), the excess mortality ranged from 7.4% to 18.5% (A1: P < .003, A2: Adjusted P < .040). Comparison of the excess mortalities’ distribution to the null counterfactual showed that the mortality increase was highly significant across Europe (Adjusted P < .001).

## Introduction

Coronavirus disease 2019 (COVID-19) is a human-to-human transmissible infectious disease that reached the size of a pandemic in early March 2020, causing more than 5.5 million official deaths worldwide in less than two years [1]. Although the damage caused by this novel coronavirus (called SARS-CoV-2) has been catastrophic, the harms in the counterfactual scenario of the absence of non-pharmaceutical measures, therapies, and vaccines would have been even more dramatic [2,3]. Besides, the official number of COVID-19 deaths is plausibly an underestimate of the real figure, given the poor testing capabilities in the initial phase [4,5]. For this reason, scientists have begun comparing mortality statistics from previous years with current ones to highlight the actual epidemiological impact of COVID-19. Nevertheless, such a comparison is far from simple. Specifically, real time series can present critical issues such as trends, seasonalities, and level shifts. Therefore, direct comparisons between pandemic data and previous year averages can be improper or even misleading. Moreover, further problems concern the trend estimation: indeed, determining the beginning of one trend and the end of another is a process that requires a high number of iterations to find the statistically most significant fit. In this regard, the Division of Cancer Control and Population Sciences of the National Institutes of Health has developed free software - called Joinpoint - to search for the best linear subtrends within a timeseries [6]. In this paper, Joinpoint was used to compare the annual mortality rates of 27 European countries before and after COVID-19 (from 2000 to 2020). The purpose of the manuscript is to provide epidemiologically relevant data and solid evidence of the COVID-19 dangerousness. Indeed, various conspiracy hypotheses have argued that a substantial number of patients died from other causes, even if tested positive to SARS-CoV-2 [7]. Since risk perception is strongly influenced by how information and graphs are presented, a simple and intuitive figure has been developed to show the results to the lay public [8]. Finally, although more detailed surveys (e.g., stratified by age groups and periods) have been conducted, a more straightforward approach can provide clearer evidence and require fewer assumptions, reducing the likelihood of interpretative errors.

## Methods

### Summary of the procedure

Annual mortality data from 2000 to 2020 for all European countries were collected from the website “The World Bank” and downloaded in “.xls” format [9]. Mortality for 2020 has been obtained from the “Eurostat” website [10]. The time series were plotted for an initial check for normality and absence of marked outliers and heteroskedasticity. After that, the “Joinpoint” software - provided by the “Division of Cancer Control & Population Science” of the National Institutes of Health (NIH) - was adopted to break the time series into linear subtrends [6]. The last subtrend found was then analyzed and confirmed by a graph check and a linear regression analysis performed with the “XLSTAT” tool for Microsoft Excel v.2112 [11]. In particular, the tool automatically quantitatively checks the assumptions of the linear regression. Finally, the residual between the model’s prediction for 2020 and the observed 2020 value was calculated for each time series. The Grubb test for high outliers was applied to verify whether the 2020 residual was out of the distribution of previous residuals [12]. One sample t-test was exploited to compare two mortality distributions: the observed one and the counterfactual centered in 0.

### Statistical details

Joinpoint regression. Most of the settings have been left at their defaults. The changed settings are specified below: Type of variable = Crude Rate (Death rate), Log transformation = No {y=xb}, Independent Variable = Year. In some cases, denoted with *, we have forced the model to introduce at least one joinpoint to fit the last values of the timeseries better (** was used for two joinpoints). All the graphs of these analyzes are reported as supplementary material to allow the reader an independent evaluation [13]. The acronym JPR-i indicates the i number of joinpoints. The absence of trend was indicated with NT.

Linear regression. Ordinary least square linear regression from the XLSTAT package was used to model the annual mortality trend from the last joinpoint through 2019. The standard assumptions of the model - i.e., normality of residuals, absence of outliers, and homoscedasticity - were automatically verified by Shapiro-Wilk and F-tests. The residual of 2020 was indicated with r_20 while the distribution of residuals from the last joinpoint up to 2019 with d. The entire analysis is available as a supplementary Excel file to allow the reader an independent evaluation.

Multi-testing adjustment. Two different approaches, called A1 and A2, were used. A1: The significance of a global test (i.e., Europe) was evaluated; then, other subtests (i.e., countries) were implemented without corrections (Siegel compromise). A2: The Holm-Bonferroni method was employed with threshold α = .05 and number of hypotheses m = 28 (A2).

P-values. A1: P-values were used as graded measures of the strength of evidence against the null hypothesis. Therefore we have not adopted dichotomic significance thresholds. However, we have divided the degrees of significance into low (P>.300), medium (.100<P≤.300), fair (.050<P≤.100), moderate (.010<P≤.050), and high (P≤.010). Indeed, we believe that this subdivision may be a compromise between the standard adoption of an a priori threshold - the purpose of which is to avoid interpretative biases - and the critical issues highlighted by Greenland et al. [14]. A2: P-values were used as graded measures of the strength of evidence against the null hypothesis. In this case, the significance was divided into high (P≤.050), moderate (.050<P≤.100), and low (P>.100).

## Results

The excess annual mortalities in 2020 of European nations (2020 Δ%) are shown in Table 1.

**Table 1.** Excess annual mortalities in 2020 in all European countries. Δ% = percentage increase, r_20 = 2020 residual, JPR-i = joinpoints number, NT = no trend, JP = joinpoint.

| Nation | Outlier | 2020 $\Delta\%$ | SE | r_20/SD | P | Adjusted P | Trend | JP Years |
| --- | --- | --- | --- | --- | --- | --- | --- | --- |
| Austria | 2020 | 10.9 | 0.5 | 5.1 | 0.001 | 0.015 | NT | NA |
| Belgium | 2020 | 14.4 | 0.6 | 8.3 | 0.000 | 0.001 | JPR1/NT* | 2007 |
| Bulgaria | 2020 | 16.2 | 2.2 | 10.8 | 0.000 | 0.000 | JPR-0 | NA |
| Croatia | 2020 | 7.0 | 6.9 | 2.9 | 0.191 | 0.999 | JPR-1* | 2013 |
| Cyprus | 2017 | 0.4 | 1.3 | 1.1 | 0.551 | 0.999 | JPR-1 | 2015 |
| Czech Rep. | 2020 | 16.4 | 0.6 | 7.7 | 0.000 | 0.000 | NT | NA |
| Denmark | 2018 | -0.4 | 2.9 | -0.4 | 0.029 | 0.352 | JPR-1 | 2014 |
| Estonia | 2020 | 1.8 | 0.4 | 1.7 | 0.766 | 0.999 | JPR-2/NT | 2007; 2011 |
| EUROPE | 2020 | 10.0 | 2.4 | 8.1 | 0.000 | 0.001 | JPR-1 | 2008 |
| Finland | 2010 | 0.5 | 1.1 | 0.7 | 0.834 | 0.999 | JPR-1 | 2005 |
| France | 2020 | 8.8 | 1.9 | 8.6 | 0.000 | 0.008 | JPR-1# | 2007 |
| Germany | 2014 | 2.5 | 2.2 | 2.0 | 0.567 | 0.999 | JPR-1 | 2007 |
| Greece | 2012 | 2.6 | 4.4 | 1.2 | 0.999 | 0.999 | JPR-1 | 2010 |
| Hungary | 2020 | 10.4 | 0.4 | 6.2 | 0.000 | 0.002 | NT | NA |
| Ireland | N.A. | 1.5 | 0.4 | 1.0 | N.A. | N.A. | JPR-2/NT | 2002; 2008 |
| Italy | 2020 | 18.5 | 3.9 | 9.2 | 0.000 | 0.000 | JPR-1* | 2006 |
| Latvia | 2019 | 2.2 | 2.4 | 1.8 | 0.682 | 0.999 | JPR-2 | 2006; 2009 |
| Lithuania | 2020 | 12.5 | 0.7 | 5.5 | 0.001 | 0.017 | JPR-1/NT | 2006 |
| Luxembourg | 2020 | 4.8 | 0.7 | 2.8 | 0.180 | 0.999 | JPR-1/NT* | 2014 |
| Malta | 2012 | 3.5 | 0.7 | 1.2 | 0.395 | 0.999 | NT | NA |
| Netherlands | 2020 | 7.8 | 2.3 | 6.9 | 0.000 | 0.006 | JPR-1 | 2009 |
| Poland | 2020 | 13.4 | 4.8 | 9.3 | 0.001 | 0.018 | JPR-1* | 2014 |
| Portugal | 2020 | 7.4 | 6.5 | 6.0 | 0.003 | 0.040 | JPR-1 | 2011 |
| Romania | 2020 | 13.2 | 2.8 | 9.2 | 0.000 | 0.000 | JPR-2** | 2007 |
| Slovak Rep. | 2020 | 10.4 | 0.4 | 6.6 | 0.000 | 0.001 | NT | NA |
| Slovenia | 2020 | 13.5 | 7.4 | 10.2 | 0.000 | 0.001 | JPR-1 | 2011 |
| Spain | 2020 | 13.0 | 5.1 | 5.9 | 0.002 | 0.025 | JPR-1 | 2010 |
| Sweden | 2020 | 8.4 | 1.9 | 6.4 | 0.000 | 0.001 | JPR-0 | NA |

The general European situation is shown in Figure 1.

**Figure 1.**
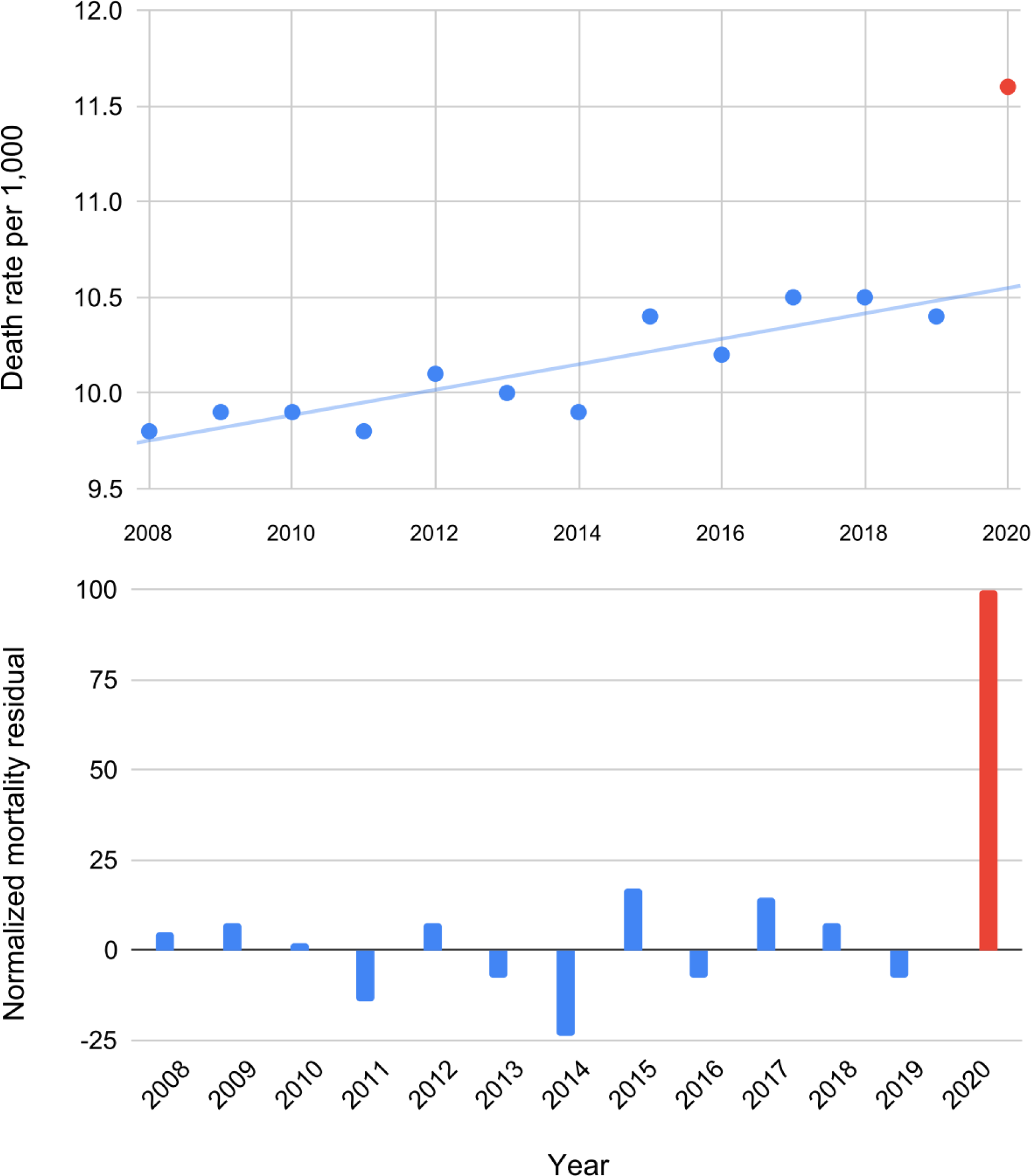
Differences between model predictions and observed death rates from 2008 to 2020 (residuals) in Europe. The abnormal increase in mortality during 2020 is visible to the naked eye.

The Grabb test identified anomalous excess mortality during 2020 in 74.1% of cases (20/27 EU countries). A generally high statistical significance was achieved in 59.3% of cases (16/27 EU countries, A1: P < .003, A2: Adjusted P ≤ .040. The percentage death rate excess ranged from 7.4% to 18.5% in the latter. However, all countries except Denmark recorded a positive death rate surplus during 2020 (mean = 8.2, SD = 5.7). Comparing this distribution with that translated into 0, we obtain t = 7.5 (df = 26, P < .001, Adjusted P < .001). Therefore, the increase in annual mortality was highly/moderately significant across the whole of Europe in 2020. Finally, the analysis revealed an increasing mortality trend in Europe over the last decade in 20 out of 27 countries (74.1%). The overall trend can be observed in Figure 1.

## Discussion

### Main findings

This brief research identified a globally highly significant and marked mortality increase in Europe during 2020. A heavy outlier was identified during 2020 compared to the model’s predictions in most countries. In particular, Austria, Belgium, Bulgaria, Czech Republic, France, Hungary, Italy, Lithuania, Netherlands, Poland, Portugal, Romania, Slovak Republic, Slovenia, Spain, and Sweden were affected by an abnormal national casualty rate. Croatia and Luxembourg also recorded a potential outlier but with less statistical significance. Furthermore, the comparison of the null counterfactual scenario (i.e., no mortality increase) with the observed one revealed a highly/moderately significant rise even for countries in which the Grabb test did not find an outlier. These findings provide strong evidence in favor of the absence of previous endogenous reasons capable of explaining the excess deaths in Europe. Nevertheless, an increasing mortality trend has characterized three out of four European nations over the past decade.

### Comparison with other literature

A vast literature discussed the virological and epidemiological reasons why COVID-19 is a dangerous disease [15]. A meta-analysis on over 423,000 patients worldwide found a pooled prevalence of mortality among hospitalized of 18%, as well as an increased mortality risk for elderly patients (pooled odds ratio and hazard ratio 2.6 and 1.3, respectively) [16]. A French study on 89,500 patients with COVID-19 and 46,000 patients with influenza compared the severity of the two diseases, highlighting that severe acute respiratory syndrome coronavirus 2 causes considerably more respiratory complications and has significantly higher mortality [17]. Also, the novel coronavirus 2019 has a higher contagion power than flu viruses [18]. Finally, there are valid molecular and biological rationales that explain the high virulence of COVID-19 [7, 19]. We conclude that COVID-19 is the main reason for the dramatic scenario depicted in this study. Similar conclusions were reached by a recent article by Dr. David Adam, which discusses the real death toll due to the new 2020 pandemic [20].

Although mortality increases with age and the infection severity is linked to a wide range of local factors and age-related comorbidities [7, 15-19], it is wrong to conclude that COVID-19 is not harmful to young people. In particular, i) long-COVID (long-term effects of COVID-19) is a condition not uncommon in the younger age groups [21], and ii) the uncontrolled spread favors the appearance of variants of concern, capable of being much more dangerous even for the youngest (e.g., Delta and Omicron) [22]. Alongside this, joinpoint regression revealed growing mortality trends in almost all of Europe. This fact should alert health authorities beyond COVID-19. In conclusion, the author of this paper wishes these results to be helpful to the scientific community to estimate the actual number of deaths due to COVID-19 and to the population to achieve an adequate risk perception. Finally, the reasons for the growing European mortality must be investigated.

### Strengths

This short paper uses an innovative method to investigate the European epidemiological scenario through two complementary procedures: i) the historical analysis of time series through joinpoint regression and ii) the search for outliers in residual distributions. In addition, two different approaches are used to interpret the statistical significance of the results to allow the reader an independent evaluation and reduce the effect of the author’s biases. Finally, Figure 1 has been conveniently constructed to be easy to interpret in order to provide an understandable reference for a lay public: specifically, simple linear scales and models have been adopted, thus making the 2020 statistical anomaly clear and evident [7].

### Limitations

This research does not examine any non-linear subtrends capable of generating false and misleading statistical anomalies [7]. Besides, the models employed cannot explore the causal nature of the results found. Finally, the differences concerning countermeasures, age, sex, comorbidities, and other local factors are not investigated. However, non-linear subtrends are rare phenomena, and it is implausible that they occurred simultaneously in over 16 countries. Furthermore, these findings acquire significance in light of the known literature on COVID-19.

## Conclusions

This paper provides clear statistical evidence on the significant impact of COVID-19 on the mortality increase recorded in Europe during 2020. Together with the virological examinations found in literature, these findings confirm the dangerousness of the novel coronavirus 2019 and definitively disclaim the conspiracy hypotheses on its harmlessness. Alongside this, the European health authorities are called upon to investigate the reasons for the growing mortality trend identified in the last decade. Finally, this manuscript shows that joinpoint regression is a powerful tool for identifying trends.

## Data Availability

All data produced in the present work are contained in the manuscript

## Acknowledgments

I thank Dr. Akshaya Srikanth Bhagavathula for introducing me to the joinpoint regression method. I also thank Dr. Lucia Castaldo for her support in writing this manuscript.

## References

1. WHO Coronavirus (COVID-19) Dashboard. (2022). Accessed: January 18, 2022: https://covid19.who.int/.

2. Mendez-Brito A, El Bcheraoui C, Pozo-Martin F: Systematic review of empirical studies comparing the effectiveness of non-pharmaceutical interventions against COVID-19. J Infect. 2021, 83(3):281-293. 10.1016/j.jinf.2021.06.018

3. Harder T, Külper-Schiek W, Reda S, et al.: Effectiveness of COVID-19 vaccines against SARS-CoV-2 infection with the Delta (B.1.617.2) variant: second interim results of a living systematic review and meta-analysis, 1 January to 25 August 2021. Euro Surveill. 2021, 26(41):2100920. 10.2807/1560-7917

4. Modi C, Böhm V, Ferraro S, et al.: Estimating COVID-19 mortality in Italy early in the COVID-19 pandemic. Nat Commun. 2021, 12(1):2729. 10.1038/s41467-021-22944-0

5. The true death toll of COVID-19 Estimating global excess mortality. (2022). Accessed: January 18, 2022: https://www.who.int/data/stories/the-true-death-toll-of-covid-19-estimating-global-excess-mortality.

6. Joinpoint Trend Analysis Software v.4.9.0.0. (2021). Accessed: August 17, 2021: https://surveillance.cancer.gov/joinpoint/.

7. Rovetta A, Bhagavathula AS: Dying From COVID-19 or With COVID-19: A Definitive Answer Through a Retrospective Analysis of Mortality in Italy [Preprint]. medRxiv. 2021, 10.1101/2021.12.22.21268212

8. Ancker JS, Senathirajah Y, Kukafka R, et al.: Design features of graphs in health risk communication: a systematic review. J Am Med Inform Assoc. 2006, 13(6):608–618. 10.1197/jamia.M2115

9. Death rate, crude (per 1,000 people). (2021). Accessed: December 29, 2021: https://data.worldbank.org/indicator/SP.DYN.CDRT.IN.

10. Population change - Demographic balance and crude rates at national level. (2021). Accessed: December 29, 2021: https://ec.europa.eu/eurostat/databrowser/view/DEMO_GINDcustom_60961/settings_1/table.

11. XLSTAT Free v.2021.1. (2021). Accessed: August 17, 2021: https://www.xlstat.com/en/.

12. RStudio v.4.1.2 software (library “outliers”). Accessed: August 1, 2021: https://www.rstudio.com/.

13. GitHub. Accessed: January 20, 2022: https://github.com/alex4lp/SupplementaryMaterial/blob/main/SupplementaryMaterial.zip.

14. Greenland S, Senn SJ, Rothman KJ, et al.: Statistical tests, P values, confidence intervals, and power: a guide to misinterpretations. Eur J Epidemiol. 2016, 31(4):337-350. 10.1007/s10654-016-0149-3

15. Hu B, Guo H, Zhou P, et al.: Characteristics of SARS-CoV-2 and COVID-19. Nat Rev Microbiol. 2021, 19(3):141-154 10.1038/s41579-020-00459-7

16. Dessie ZG, Zewotir T: Mortality-related risk factors of COVID-19: a systematic review and meta-analysis of 42 studies and 423,117 patients. BMC Infect Dis. 2021, 21(1):855. 10.1186/s12879-021-06536-3

17. Piroth L, Cottenet J, Mariet AS, et al.: Comparison of the characteristics, morbidity, and mortality of COVID-19 and seasonal influenza: a nationwide, population-based retrospective cohort study. Lancet Respir Med. 2021, 9(3):251-259. 10.1016/S2213-2600(20)30527-0

18. What is the difference between Influenza (Flu) and COVID-19?. (2022). Accessed: February 1, 2022: https://www.cdc.gov/flu/symptoms/flu-vs-covid19.htm.

19. Zhang Q, Xiang R, Huo S, et al.: Molecular mechanism of interaction between SARS-CoV-2 and host cells and interventional therapy. Signal Transduct Target Ther. 2021, 6(1):233. 10.1038/s41392-021-00653-w

20. Adam D: The pandemic’s true death toll: millions more than official counts. Nature. 2022, 601(7893):312–315 10.1038/d41586-022-00104-8

21. Behnood SA, Shafran R, Bennett SD, et al.: Persistent symptoms following SARS-CoV-2 infection amongst children and young people: A meta-analysis of controlled and uncontrolled studies. J Infect. 2021, S0163-4453(21)00555-7. 10.1016/j.jinf.2021.11.011

22. Funk AL, Florin TA, Kuppermann N, et al.: Outcomes of SARS-CoV-2-Positive Youths Tested in Emergency Departments: The Global PERN-COVID-19 Study. JAMA Netw Open. 2022, 5(1):e2142322. 10.1001/jamanetworkopen.2021.42322

